# Omicron infection induces low-level, narrow-range SARS-CoV-2 neutralizing activity

**DOI:** 10.1101/2022.05.02.22274436

**Authors:** Priscilla Turelli, María-Eugenia Zaballa, Charlène Raclot, Craig Fenwick, Laurent Kaiser, Isabella Eckerle, Giuseppe Pantaleo, Idris Guessous, Silvia Stringhini, Didier Trono

## Abstract

**Background:** The rapid worldwide spread of the mildly pathogenic SARS-CoV-2 Omicron variant has led to the suggestion that it will induce levels of collective immunity that will help putting an end to the COVID19 pandemics.

**Methods:** Convalescent serums from non-hospitalized individuals previously infected with Alpha, Delta or Omicron BA.1 SARS-CoV-2 or subjected to a full mRNA vaccine regimen were evaluated for their ability to neutralize a broad panel of SARS-CoV-2 variants.

**Findings:** Prior vaccination or infection with the Alpha or to a lesser extent Delta strains conferred robust neutralizing titers against most variants, albeit more weakly against Beta and even more Omicron. In contrast, Omicron convalescent serums only displayed low level of neutralization activity against the cognate virus and were unable to neutralize other SARS-CoV-2 variants.

**Interpretation:** Moderately symptomatic Omicron infection is only poorly immunogenic and does not represent a substitute for vaccination.

**Funding:** EPFL COVID Fund; private foundation advised by CARIGEST SA; Private Foundation of the Geneva University Hospitals; General Directorate of Health of the canton of Geneva, the Swiss Federal Office of Public Health.

---

Omicron (also referred to as BA.1 or B.1.1.529) was identified as a new SARS-CoV-2 (Severe Acute Respiratory Syndrome Coronavirus 2) variant in late 2021 and rapidly became dominant so as to account today for a large majority of new infections worldwide^1^. Omicron differs from previously prevalent SARS-CoV-2 variants at more than thirty amino acid positions within Spike, the protein responsible for mediating viral entry through recognition of the ACE2 receptor and targeted by SARS-CoV-2 neutralizing antibodies. Accordingly, Omicron escapes neutralization by most monoclonal antibodies available for the treatment of COVID19 or by pre-Omicron convalescent sera and can readily cause infections in individuals who have been either previously infected with other SARS-CoV-2 variants or subjected to full vaccination regimens, all current vaccines being derived from the original SARS-CoV-2 strain or from close relatives^2–7^.

Likely due to a combination of these immunological features and of variant-specific virological properties, Omicron is highly contagious yet generally causes milder symptoms compared with other SARS-CoV-2 variants including its immediate predecessor Delta^8,9^. This has led to the hope that the sweeping propagation of Omicron in the world population might contribute to put an end to the COVID19 pandemics by inducing, at a low cost for global health, robust levels of protective immunity including in non-vaccinated populations.

As a step towards probing this hypothesis, we measured levels of variant-specific neutralizing antibodies in the serum of a group of 23 individuals recently infected with Omicron without reported prior exposure to or vaccination against SARS-CoV-2, and whose infection did not require hospitalization (table 1). Serum samples from these individuals were collected between 3 and 74 days after a positive PCR test (with a median time of 39 days). Age-matched groups of individuals previously not infected but vaccinated with 2 doses of either Moderna or Pfizer vaccines (n=8 for each group) or infected with Alpha (B.1.1.7, n=10) or Delta (B.1.617.2, n=13) variants were used as a comparison with distribution of participants according to age, sex and time post infection or vaccination indicated in figure 1A-C. All vaccinated individuals tested positive for anti-S antibodies only while Alpha and Delta convalescents all tested positive for both anti-N and anti-S antibodies (figure 1D,E). Out of 23 Omicron convalescent sera, 17 and 20 were positive for anti-S and anti-N antibodies, respectively, albeit to a low level, strongly suggesting a lower global humoral immune response after infection by this compared with other variants.

**Figure 1:**
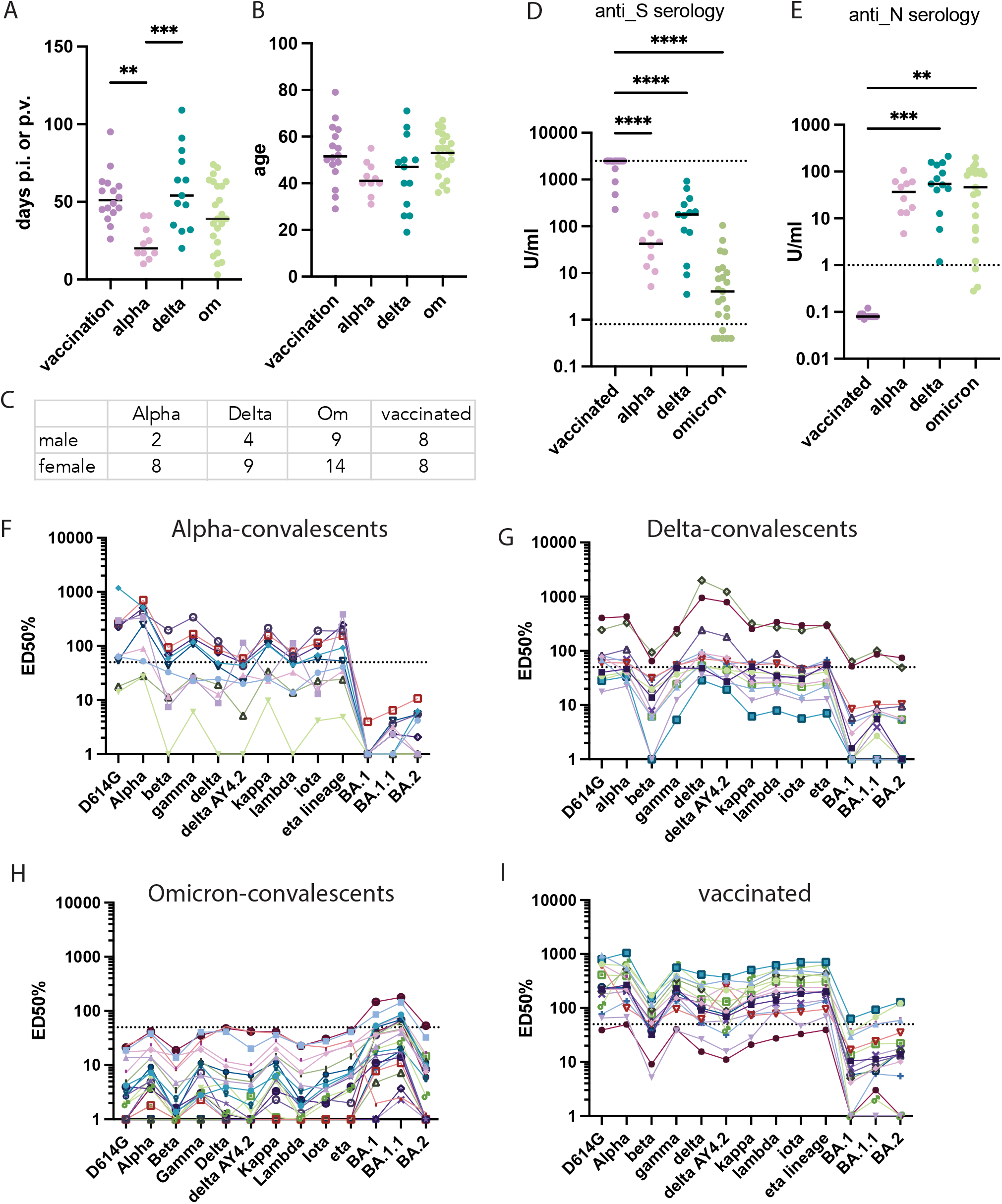
Omicron infection provides neither a strong nor a broad humoral response. (A) Sera from people who either received 2 doses of mRNA vaccines (n=16) or were infected with alpha (n=10), delta (n=13) or Omicron strains only (n=15) were collected at the indicated time post infection or vaccination. Age (B) and sex (C) distribution of volunteers. (D): Roche anti-S serology of the participants at the collection time. Values <=0.4 and values >=2500U/ml are plotted at 0.4 and 2500 (upper dashed line) respectively. The positivity threshold is fixed at 0.8U/ml (lower dashed line) (E): Roche anti-N serology. Lower limit for positivity is fixed at 1 (dashed line). a to e: The median value is plotted in each scatter dot plot. F to I: Neutralizing activities of sera from each indicated group were assessed for their ability to neutralize D614G-ancestral or indicated Spike variants in the multiplex S3-ACE2 cell-free assay. The mean serum dilution needed to achieve 50% neutralization (ED50%) is indicated for each serum and variant. Sera with an ED50% > 50 (dashed line) are considered neutralizing^10^. Sera with ED50% <=1 are plotted at 1. Only significant p-values are represented with **p<0.01, ***p<0.001, ****p<0.0001.

To measure variant-specific neutralizing antibodies, we first used a cell-free surrogate Spike-ACE2 binding interference assay, the results of which we previously demonstrated correlate faithfully with those of cell-based live virus neutralization systems^10^. We used trimeric Spike proteins derived from past variants and currently circulating lineages or sublineages including the most recent Omicron BA.1, BA.2 and BA.1.1 strains. The results revealed that individuals previously infected with the Alpha variant, except for one, developed antibodies capable of neutralizing not only the cognate Spike protein and its D614G predecessor, but also those from several more recent variants such as Gamma, Kappa and Eta, albeit with slightly decreased efficiency against Beta, Delta, Delta AY4.2, Lambda and Iota. However, Alpha-convalescent serums displayed on average a 2-log reduction in neutralization titers against all three Omicron sublineages (figure 1F). Sera harvested after Delta infection could efficiently neutralize most Spike variants including the Delta AY4.2 sub-lineage bearing the T95I, Y145H and A222V mutations, but were much less effective against Beta and even more so against the three Omicron subvariants (figure 1G). After Omicron infection, SARS-CoV-2 neutralizing titers were uniformly lower and displayed no significant activity against any of the ten other Spike variants tested through this assay. Further and concerningly, only 4 and 9 out of 23 individuals displayed titers around the 1:50 ED50 cut-off against BA.1 and its BA.1.1 sublineage, respectively, with for all an additional major reduction in BA.2 Spike neutralization (figure 1H). Finally, sera from mRNA-vaccinated individuals were strongly neutralizing against most variants but were significantly less potent against Beta and even more markedly against the 3 Omicron sublineages (Figure 1h). Noteworthy, in none of these subgroups of individuals were anti-S serological titers predictive of variant-neutralizing activity (appendix p 1).

To consolidate further these data, we used a cell-based neutralisation assay with Omicron BA.1 live virus (figure 2A). None of the Alpha-convalescent sera was able to neutralize this variant, even at the starting 1:10 dilution. Some prevention of virus-induced cytopathic effect (CPE) was obstained with the 2 Delta-convalescent sera (D-1 and D-2) found to have the highest neutralization titers against Omicron Spike in the cell-free assay (ED50%=64 and 52 respectively). Finally, low levels of Omicron-neutralizing activity (serum dilution 1:10 to 1:30 max.) were detected in 17 of the 23 Omicron-convalescent individuals, the same previously identified as displaying an ED50%>=7.8 in our cell-free assay. We extended this analysis by selecting the 5 serums from previously Omicron-infected individuals with the highest anti-S titers and probing their ability to neutralize live ancestral D614G, Alpha, Beta, Gamma, Delta and Omicron BA.2 viruses (figure 2B). Some neutralization of the BA.2 strain was clearly detected for all but 1 of these serums. Only 2 serums (Om-14 and Om-18) showed detectable neutralizing activity on other strains (D614G, alpha and Delta viruses). None of the 3 other serums could block any of the isolates, while all isolates could be neutralized by serum from a vaccinated individual used in parallel (figure 2C).

**Figure 2:**
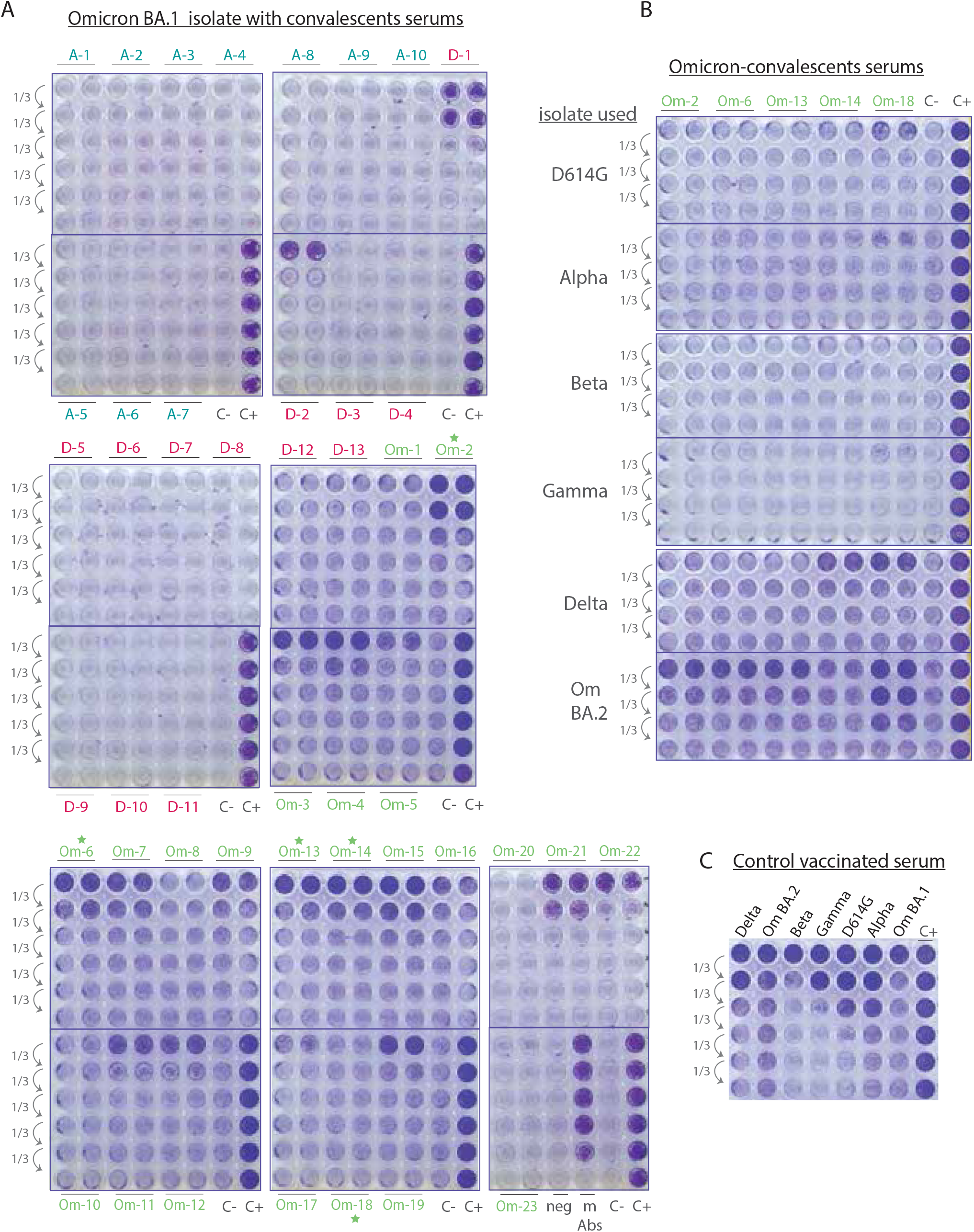
Variants convalescent serums do not neutralize BA.1 virus. (A) Neutralization potency of Alpha- (A1-10) Delta- (D1-13) or Omicron- (Om-1-23) convalescents sera was tested with 1:3 serial dilutions, in duplicates, in a CPE-based neutralization assay using Omicron BA.1 replicative isolate. (B) Same as in (A) using the indicated 5 omicron-convalescents serums which presented the highest anti-S antibodies titers and the indicated diverse isolates. (C) serum from a vaccinated individual was used as control and probed in parallel with the different isolates. C-: control wells with lysed cells incubated with virus only. C+: control cells incubated with medium only. neg: serum from pre-pandemic healthy donor. mAbs: 1:3 dilutions of a potent neutralizing monoclonal antibody used as positive control for neutralization.

In sum, these data indicate that Omicron infection generally induces on its own only low levels of neutralizing antibodies in immunocompetent individuals, and that these antibodies further display a narrow range of activity. The former may partly be related to the usual mildness of Omicron-induced symptoms and possibly to its replication in less immunogenic sites, and the latter to antigenic features differing markedly between the Spike proteins of Omicron and that of other SARS-CoV-2 variants^8,11^. Critically, our results indicate that the widespread propagation of Omicron is unlikely to induce the levels of protective collective immunity needed to curb the COVID19 pandemics, and should serve as a warning against using Omicron exposure as a surrogate for vaccination.

## Research in context

Serum neutralizing antibody titers are considered as the most significant biomarker of protective immunity against SARS-CoV-2 infection^12^. To our knowledge, only one recent peer-reviewed study has examined levels of neutralizing activity induced by Omicron infection only, but this study was limited to 20 hospitalized patients, that is, belonging to the small minority of Omicron-infected individuals presenting with serious disease hence more likely to mount a robust immune response^13^. In contrast, our analysis was restricted to individuals whose symptoms did not require hospitalization, that is, much more representative of a typical course of Omicron infection.

### Limitations of our study

No serum was available from individuals previously infected with the Omicron BA.2 subvariant, which only recently emerged in our region. Although we did not have the sequence of the infecting virus, all serums were harvested at times during which the Alpha, Delta and Omicron BA.1 variants, respectively, overwhelmingly predominated in the local population (>95%, see Methods). Convalescent serums were harvested on average 20, 54 and 39 days post Alpha, Delta and Omicron infections respectively, so that we cannot exclude that neutralizing titers had not yet reached their peak in all cases. However, previous studies indicate that antibody responses are already robust three weeks after SARS-CoV-2 infection^14,15^, which suggests that the the low titers in neutralizing antibodies of serums from our Omicron-convalescent group can not be inferred to collection time. Finally, our sample did not include individuals whose symptoms had required hospitalization, which may explain why their Omicron-directed neutralization response was weaker than recently reported for a small group of hospitalized patients^13^.

## METHODS

### Recruitment

Serum samples were obtained from participants of the Specchio-COVID19 cohort. Participants in this cohort have been recruited through one of the COVID19 serosurveys conducted in the canton of Geneva, Switzerland, and were re-invited for a follow-up serology test between April 2021 and February 2022^16^. All participants gave written informed consent and completed a questionnaire reporting on vaccination status and previous confirmed SARS-CoV-2 infections at the moment of providing a venous blood sample. The Geneva Cantonal Commission for Research Ethics approved this study (Project ID 2020-00881).

Data on type of vaccine, dates of vaccination, and date of PCR positive test were self-reported by the participant at the time of blood drawing for all samples. Reported PCR positive test dates were verified using a state centralized registry compiling SARS-CoV-2 infections or against the information provided by the participant through regular questionnaires filled during the follow-up study^16,17^.

Variant of infection was assigned based on the reported date of PCR positive test: variant of infection was assumed to be Alpha if the date of the reported infection was between March 2021 and May 2021; Delta if the infection occurred between July 2021 and September 2021; or Omicron if happening between January 2022 and February 2022. For all three chosen periods the respective variant was dominant in the canton of Geneva, responsible of the vast majority of detected infections (https://www.hug.ch/laboratoire-virologie/surveillance-variants-sars-cov-2-geneve-national). For all participants, the positive PCR taken as evidence of prior infection with Alpha, Delta or Omicron SARS-CoV-2 was the only one reported.

Accordingly, none of the participants was seropositive at any of their visits within our study before this positive PCR test. Available COVID19-related data for all participants included in this analysis is presented in table 1.

### Immunoassays

Anti-SARS-CoV-2 antibodies in fresh serum samples were tested upon collection using two commercially-available immunoassays: the Roche Elecsys® anti-SARS-CoV-2 S immunoassay (Roche Diagnostics, Rotkreuz, Switzerland), detecting immunoglobulins (IgG/A/M) against the receptor binding domain of the virus spike (S) protein (#09 289 275 190, Roche anti-S); and the Roche Elecsys® anti-SARS-CoV-2 N immunoassay, detecting immunoglobulins (IgG/A/M) targeting the virus nucleocapsid (N) protein (#09 203 079 190, Roche anti-N). Seropositivity was defined using the manufacturer’s provided cut-off values: titer ≥ 0.8 U/mL for the Roche anti-S and index ≥ 1.0 for the Roche anti-N immunoassays. All samples were then frozen and stored at -20°C.

### Spike proteins production and purification

Mutations cloned on SARS-Cov2 Spike variants and found on viral isolates are listed in table 2. Production of nCov2019(D614G), Alpha, Beta and P.1 variants has already been described^18^. Mutations found in other variants have either been cloned by PCR-mediated mutagenesis in the Wuhan codon-optimized ORF (Delta AY.4.2, Iota, Eta, Omicron BA.1.1), gene synthesis (Delta, Kappa, Lambda) or gBlocks assembly (Omicron BA.1). RNA isolated from an anonymized leftover sample of an individual confirmed to be SARS-CoV-2 Omicron BA.2 strain infected was reverse transcribed into cDNA. The Omicron Spike ectodomain was amplified by PCR and introduced by In-Fusion cloning into the nCoV-2P-F3CH2S plasmid, replacing the original wild-type Spike^19^. The 2 prolines (P986-P987) and the furin cleavage site mutations (residues 682-685 mutated to GSAS) stabilizing the Spike protein in the trimeric prefusion state were further introduced simultaneously by PCR and In-Fusion-mediated site directed mutagenesis as previously described^10^, and the full Omicron BA.2 ORF was sequence verified. All the final constructs encode the Spike ectodomains, containing a native signal peptide, the 2P and furin cleavage site mutations, a C-terminal T4 foldon fusion domain to stabilize the trimer complex followed by C-terminal 8x His and 2x Strep tags for affinity purification. The trimeric Spike variants were produced and purified as previously described^10^.

### S^3^-cell free neutralization assay

Trimeric-Spike variants production and purification and Spike protein beads coupling were performed as previously described^10^. Neutralization assays were done in 96-well plates as previously described^10^, with 10 multiplexed Spike variants and 8 μl of serum per well for the starting dilution. Briefly, 1:3 serial dilutions of sera were incubated 1 hour with the Spikes before ACE2 mouse Fc fusion protein (produced and purified as previously described^10^) was added and binding detected with an anti-mouse IgG-PE secondary antibody (OneLambda, Thermo Fisher Scientific). Control wells were included on each 96-well plate with each variant Spike-coupled beads alone. Mean fluorescence intensity (MFI) for each of the beads alone without serum were averaged and used as the 100% binding signal for the ACE2 receptor to the bead coupled spike trimer. MFI obtained for D614G Spike using a high concentration of imdevimab (RGN10987, 1 μg/ml) known to neutralize the ancestral strain was used as the maximum inhibition signal^20^. The percent blocking of the Spike protein trimer-ACE2 interaction was calculated using the formula: % Inhibition = (1- ([MFI Test dilution – MFI Max inhibition] / [MFI Max binding – MFI Max inhibition]) × 100). Serum dilution response inhibition curves were generated using GraphPad Prism 8.3.0 NonLinear four parameter curve fitting analysis of the agonist versus response, and ED50% values extracted using an in-house script.

### SARS-CoV-2 live virus stocks

All the biosafety level 3 procedures were approved by the Swiss Federal Office of Public Health (authorization A202952/3). The SARS-CoV2 D614G (EPI_ISL_414019), Alpha (EPI_ISL_2131446), Beta (EPI_ISL_981782), Gamma (EPI_ISL_981707), Delta (EPI_ISL_1811202), Omicron BA.1 (EPI_ISL_7605546) and Omicron BA.2 (EPI_ISL_8680372) early passage isolates were obtained and sequenced at the Geneva University Hospitals. Viral stocks were prepared with the early isolates in EPISERF on VeroE6 or Calu-3 cells, aliquoted, frozen and titrated on VeroE6 cells.

### SARS-CoV-2 live virus cell based cytopathic effect neutralization assay

Neutralization assay was performed as previously described^18^ except that serial sera dilutions were done in EPISERF instead of DMEM 2% FCS. In brief, 3000 plaque forming units per well of viral isolate were pre-incubated for 1 hour @ 37°C with 1:3 serial dilutions of sera prepared in duplicates in 96-well plates. The mix was then applied on VeroE6 cells and cytopathic effect detected 48hours later with Crystal violet staining.

## Data Availability

All data produced in the present study are available upon reasonable request to the authors

## ACKNOWLEDGEMENTS

We thank Florence Pojer and the Protein production and structure Core facility at EPFL for the Spike variants production, Meriem Bekliz and the Virology laboratory of Geneva University Hospital for the Omicron RNA samples and variant isolates collection, Julien Duc for the development of in-house scripts to generate neutralization curves, Géraldine Poulain and Pierre Lescuyer for sample preparation, Isabelle Arm-Vernez for serological studies, Julien Lamour and Jennifer Villers for help with cohort data analyses. We are grateful to all members of the Specchio-COVID19 team and all participants for their precious contribution to this study.

## SUPPLEMENTARY FIGURE LEGENDS

**Figure S1:**
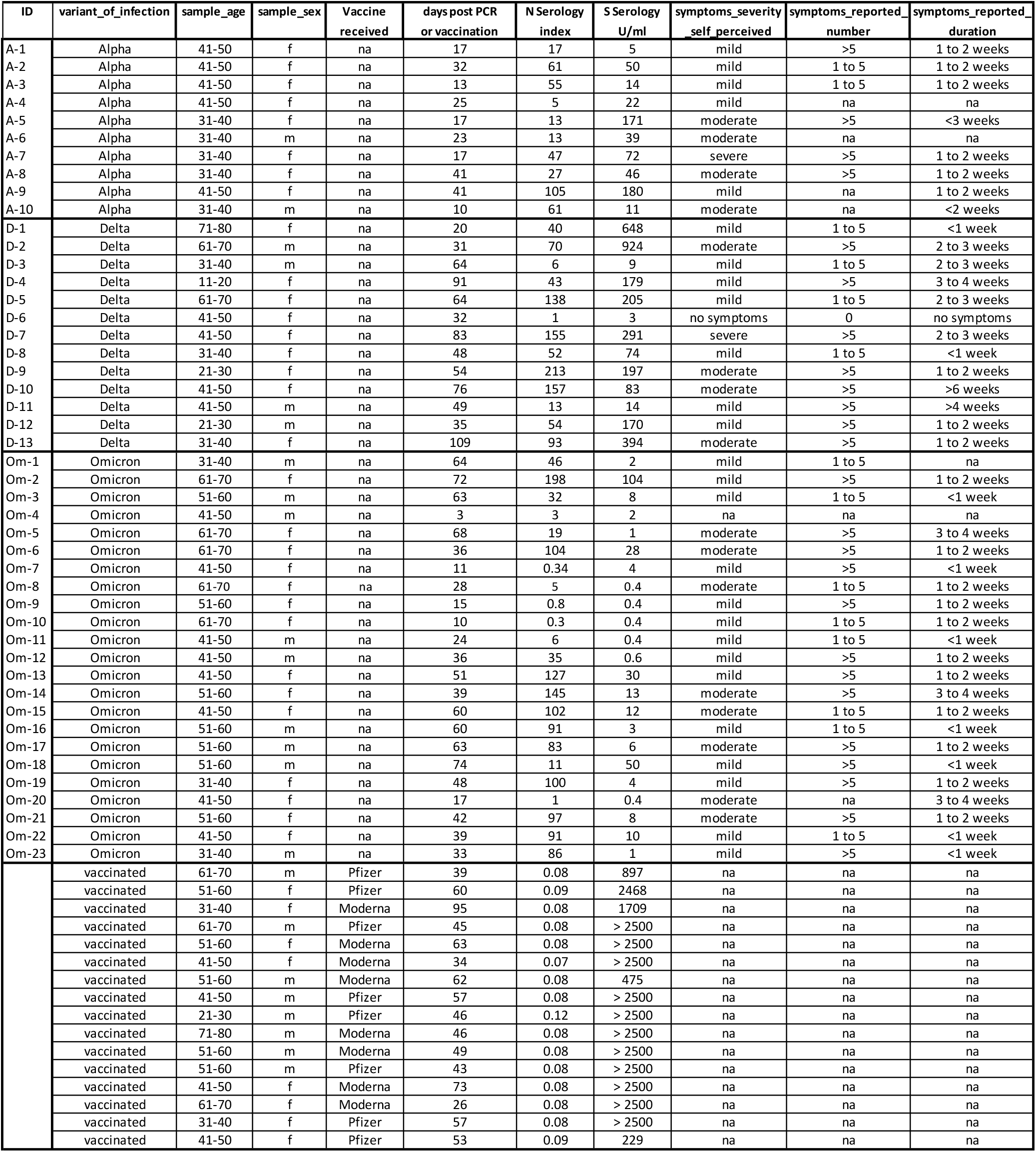
Anti-S serology is poorly correlated to neutralizing antibodies titers (A)Roche anti-S serology titers of Alpha, Delta or Omicron convalescents were plotted versus ED50% found for Alpha, Delta and Omicron Spikes respectively. Anti-S serology of vaccinated people was plotted versus ED50% for each of the 3 Spike variants. Values <=0.4 and values >=2500U/ml are plotted at 0.4 and 2500 respectively. ED50% <=1 are plotted at 1. The positivity thresholds are fixed at 0.8U/ml and 50 for S-serology and ED50% respectively (dashed lines). (B)ED50% found for Alpha, Delta and Omicron Spikes versus days post infection or vaccination of the participants. The threshold for positivity is at dilution 50 for ED50% and is indicated with a dashed line.

**Figure S2:**
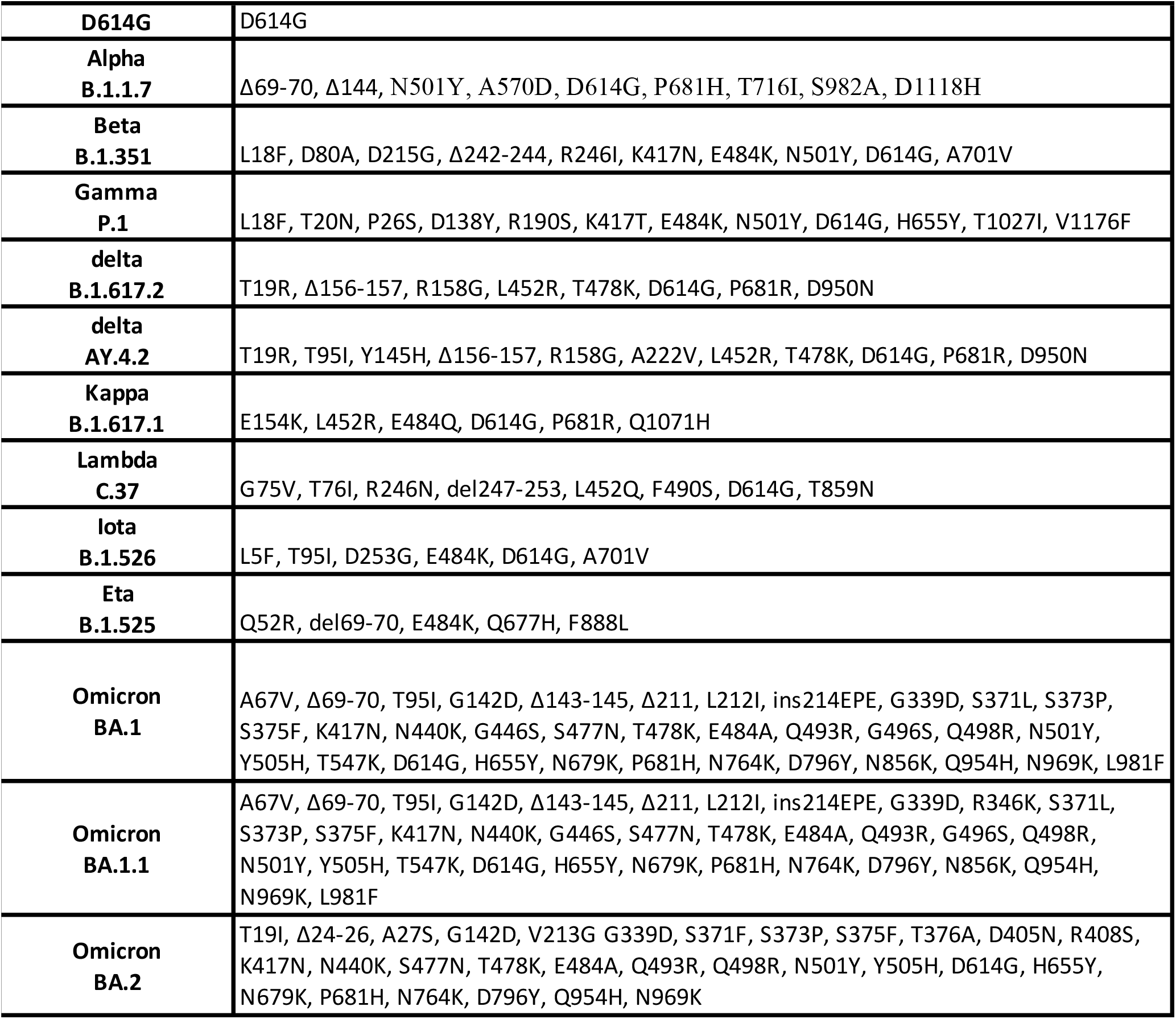
Prior infection with the Alpha or Delta strains confers neutralizing titers against homologous variants (A) Serums from individuals previously infected with Alpha viruses were used in a CPE-based neutralization assay using and Alpha replicative isolate. (B) same as (A) but with serums from individuals previously infected with a Delta virus and probed with a Delta replicative isolate. C-: control cells incubated with virus only. C+: control cells incubated with medium only. Ø: empty wells.

