## Supplementary figures and images for "Omicron infection induces low-level, narrow-range SARS-CoV-2 neutralizing activity"

### Supplemental Figure 1

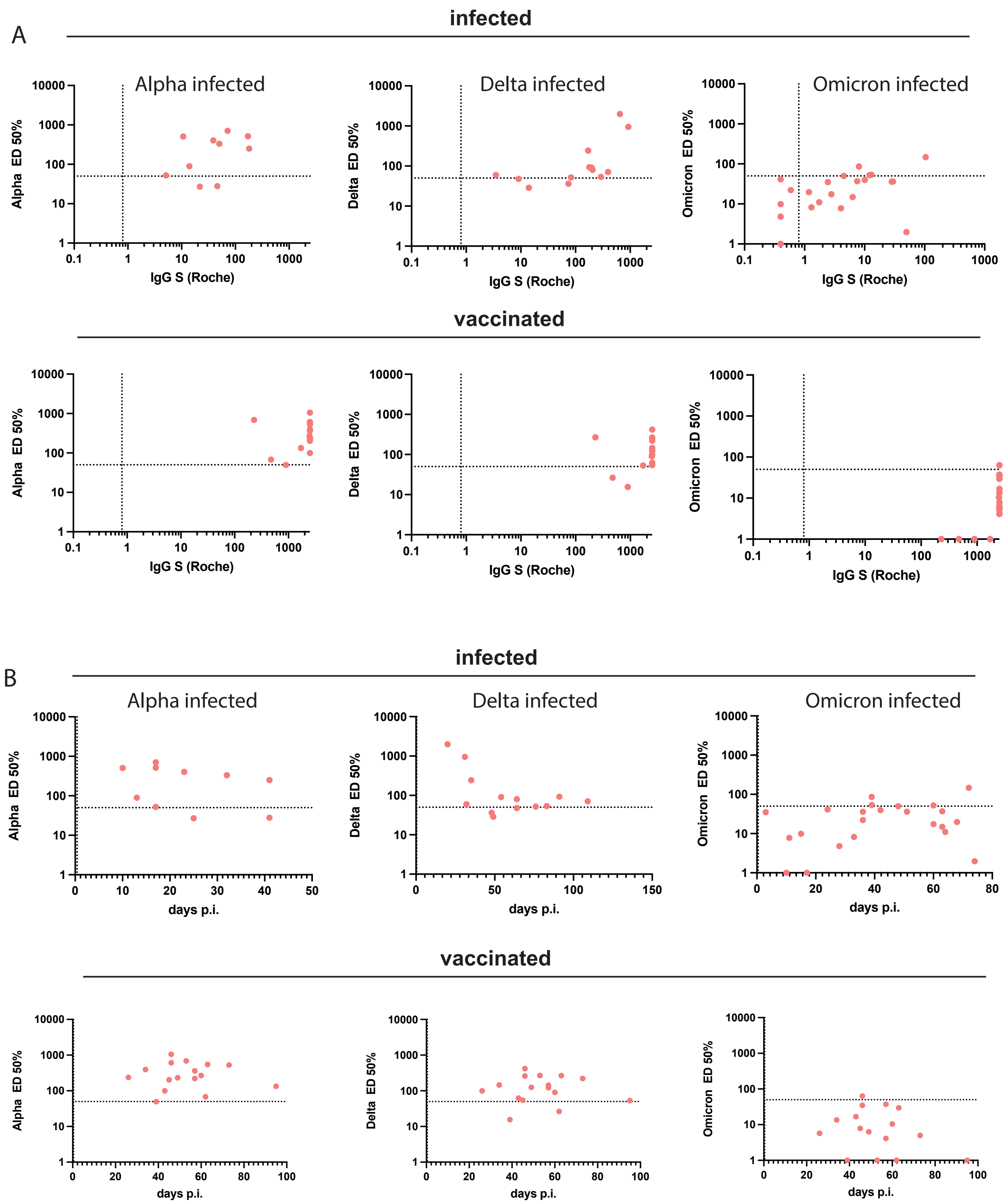

### Supplemental Figure 2

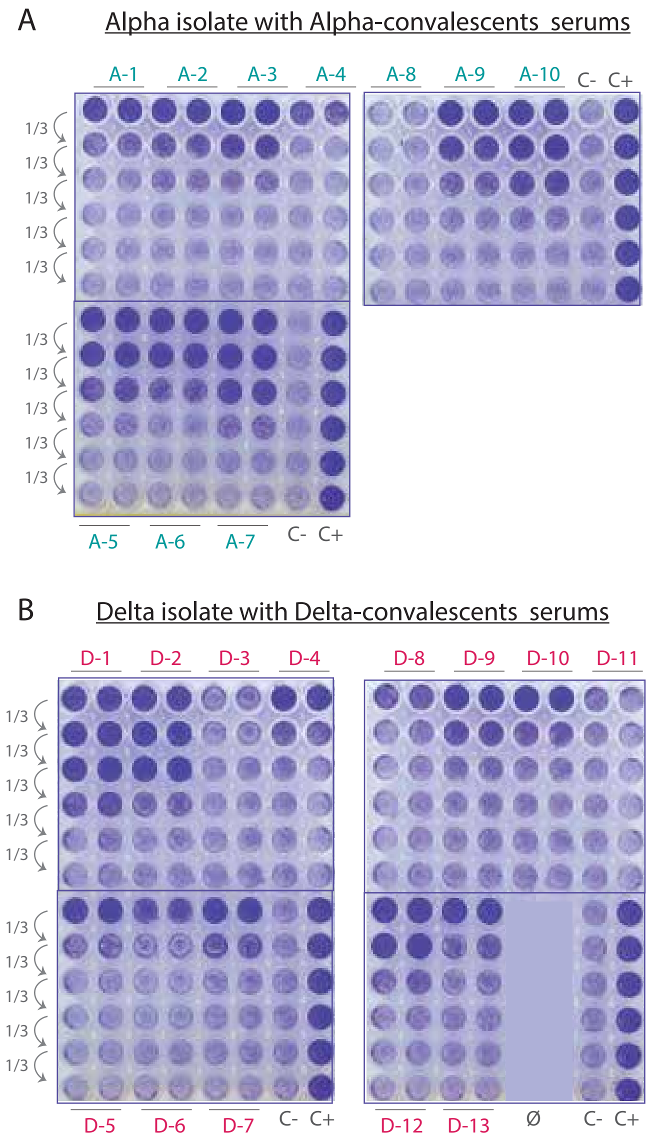
